# High 3-hydroxybutyrate concentrations in the placenta-produced amniotic fluid in the human uterus

**DOI:** 10.1101/2023.08.09.23293873

**Authors:** Takumi Satoh, Takeo Shibata, Emi Takata, Masahiro Takakura, Jia Han, Sohsuke Yamada

## Abstract

In this study, we report first high concentrations of a ketone body, 3-hydroxybutyrate (3HB) in the amniotic fluid in humans. Although 3HB concentrations in the maternal blood are approximately 0.1, those in the amniotic fluid are approximately 0.6 mM. High placental 3HB production is potentially key for producing and maintaining high 3HB levels in the amniotic fluid. The rate-limiting enzyme, mitochondrial 3-hydroxy-3-methylglutaryl-CoA synthase 2 (HMGCS2), is highly expressed in the cells of the chorionic plate and responsible for 3HB production. Therefore, high HMGCS2 expression maintenance is supposed to be pivotal for maintaining the 3HB supply for the human fetus. Here, we propose that humans display two pathways, an amniotic fluid- and another umbilical vein-mediated, for supplying 3HB to the human fetus. These supply pathways are supposedly essential for human brain development during the late phase of pregnancy.

**Graphical abstract:** Human fetuses are supported by 3HB from the amniotic fluid for their brain development.

**HIGHLIGTS:** 3-Hydroxybutyrate concentrations are high in the amniotic fluid in the human uterus.

The chorionic plate of the placenta highly expresses 3-hydroxy-3-methylglutaryl-CoA synthase 2.

Human fetuses may be supplied with 3HB for brain development through the amniotic fluid.

## Introduction

Ketone bodies, composed of 3-hydroxybutyrate (3HB), acetoacetate, and acetone, are fatty acid-derived energy substrates with supposed roles during starvation [1-2]. 3HB is produced through fatty acid oxidative metabolism in the mitochondria and used as an energy substrate for mitochondrial metabolism [3,4]. 3HB serves energy production in the mitochondria, affecting human health in various ways. 3HB allows for higher ATP production than glucose as 3HB could be fully oxidized [3-4]. In addition, 3HB induces various positive effects in humans [5-9].

Glucose and a ketone body (3hydroxybutyrate: 3HB) uptakes of the brain have been measured 48 years ago, taking samples from an aborted human fetus (28 weeks of gestation) and culturing for several hours at the organ level. 3HB and glucose accounted for 60% and 40%of the energy, respectively, providing direct evidence of the human fetal brain preferentially using 3HB as an energy source [10]. The human brain can rapidly grow using an energy supply of a “main-3HB and sub-glucose” system. At least, up to late pregnancy, the human brain could use 3HB as a fuel [10,11]

Certain cells of endoderm origin, such as those of the gut, liver, and placenta, exhibit high HMGCS2 expression, continuously producing 3HB both at the luminal and tissue sides [12-17]. For example, gut epithelial cells, with high HMGCS2 expression, could activate stem cell population expansion by releasing 3HB into the tissue side of the ring structure [12,13]. In particular, HMGCS2 is reportedly involved in anti-tumor effects in colorectal cancer [14,15]. Therefore, we suspect that a distinctive HMGCS2-related 3HB production system might exist in the human placenta [16,17]. We also suppose that several 3HB supplying systems could be installed in the human placenta for supplying 3HB to the human fetus.

3HB concentrations in the human and rat fetus are reportedly significantly high compared with those in adults [10,11,18]. The human fetus certainly depends on 3HB as a main energy source. One possible pathway is villi trophoblast→ umbilical vein → fetus, proposed by Muneta et al (2016) [11]. However, we suspected the existence of an alternative pathway to supply 3HB to the fetus.

## Methods and materials

### Participants and 3HB measurements

We involved nine healthy pregnant women in the study who had a cesarean section (C-section) in full-term, without any abnormal glucose tolerance or gestational diabetes mellitus. 3HB levels were measured in the umbilical vein, umbilical artery, amniotic fluid, and maternal blood. Maternal blood was drawn the day before the C-section. Umbilical venous blood and umbilical arterial blood were collected at C-section. The whole blood samples of maternal blood, umbilical venous blood, and umbilical arterial were used for the 3HB measurements. We quickly collected the amniotic fluid after the artificial rupture of the membrane at the time of the C-section with as little maternal blood as possible. The amniotic fluid was centrifuged at 3000 G for 20 minutes to remove fetal and maternal cells and blood, and the supernatant was used for 3HB measurements using Precision Exceed III (Abbott).

### Histopathology and immunohistochemistry

Histopathology and immunohistochemistry were performed as described previously[19,20]. The paraffin-embedded human placenta samples were provided by the Kanazawa Medical University (Ishikawa, Japan). The paraffin-embedded dissected placenta samples for histological examinations were cut systematically in sequential 4-μm-thick longitudinal sections using a sliding microtome (Leica SM2010R, Leica Microsystems, Wetzler, Germany). For the histological analyses of the large intestine, we captured and merged images of hematoxylin and eosin or HMGCS2 immunohistochemistry sections using the NanoZoomer Digital Pathology Virtual Slide Viewer software program (Hamamatsu Photonics Corp., Hamamatsu, Japan).

HMGCS2 immunohistochemistry staining was performed using an HMGCS2 mouse monoclonal antibody (SANTA CRUZ, sc-393256). The procedure was as follows: (1) deparaffinization and rehydration; (2) 0.5%H_2_O_2_ blocking for 10 minutes at room temperature; (3) heating (95^°^)C: in an epitope retrieval buffer (pH6.0, with citric acid) for 10 minutes then cooling down for 30 minutes at room temperature; (4) primary antibody incubation overnight at 4^∘^C (dilution: 1:500); (5) secondary antibody (Histofine Simple Stain MAX-PO424152) staining for 30 minutes at room temperature; (6) 3,3’diaminobenzidine (Histofine Simple Stain SAB-PO425011) imaging and hematoxylin counterstaining. For the HMGCS2 immunohistochemistry, we used human liver cells as a positive control.

### Statistical analyses

The data are represented as the mean ± standard deviation. For statistical analyses, we calculated the P-values according to the Mann–Whitney U test to compare the two groups.

### Ethical considerations

The present study was conducted in compliance with the ethical principles of the Declaration of Helsinki (Note of Clarification added at the 2004 World Medical Association General Assembly in Tokyo), Japan’s Act on the Protection of Personal Information, and with reference to the Ministerial Ordinance on Good Clinical Practice for Drugs (Ordinance of Ministry of Health and Welfare No.28 of March 27, 1997) and the Ethical Guidelines for Epidemiological Research established by Japan’s Ministry of Health, Labour and Welfare, and Ministry of Education, Culture, Sports, Science and Technology. Concerning the 3HB measurements, this study was approved by the Institutional Review Board at the Kanazawa Medical University (IRB #I747) and written informed consent was obtained from all study participants. In the case of the HMGCS2 immunohistochemistry, our institution did not require any ethical approval for reporting individual cases or case series.

## Results

### Participant characteristics

The average age of the nine participants enrolled in this study was 37.3 ± 4.2 years. The average week of C-section was 37.1 ± 0.3. The indications for the C-section were repeated cesarean birth (seven patients), prior myomectomies (one patient), and breech presentation (one patient). Not all pregnant women were obese or underweight (body mass index [BMI]: 25.3 ± 3.0). No patients had any pregnancy-related complications (Table 1).

**Table 1.** Patients Characteristics.

| Patients | Gravidity | Parity | Week of Delivery | Height (cm) | Body Weight (Kg) | BMI | Indication of C-section | Mode of Conception | Complications of Pregnancy |
| --- | --- | --- | --- | --- | --- | --- | --- | --- | --- |
| Patient 1 | 2 | 1 | 37 | 160.5 | 58.5 | 22.7 | Repeat Cesarean Birth | Natural Pregnancy | None |
| Patient 2 | 2 | 0 | 38 | 162 | 71.9 | 27.4 | Prior Myomectomies | Natural Pregnancy | None |
| Patient 3 | 2 | 1 | 37 | 160.1 | 62.8 | 24.5 | Repeat Cesarean Birth | Natural Pregnancy | None |
| Patient 4 | 2 | 1 | 37 | 157 | 71.4 | 29 | Repeat Cesarean Birth | Natural Pregnancy | None |
| Patient 5 | 4 | 1 | 37 | 169 | 70.6 | 24.7 | Repeat Cesarean Birth | Natural Pregnancy | None |
| Patient 6 | 3 | 2 | 37 | 153 | 70.1 | 29.9 | Repeat Cesarean Birth | Natural Pregnancy | None |
| Patient 7 | 4 | 3 | 37 | 161 | 53.6 | 20.7 | Repeat Cesarean Birth | Natural Pregnancy | None |
| Patient 8 | 4 | 2 | 37 | 166 | 67.7 | 24.6 | Repeat Cesarean Birth | Natural Pregnancy | None |
| Patient 9 | 2 | 1 | 37 | 162 | 64 | 24.4 | Breech Presentation | In vitro fertilization | None |

### HB measurements

We measured the 3HB concentrations in the amniotic fluid, umbilical vein, umbilical artery, and maternal blood (Figure 1). The 3HB concentrations in the amniotic fluid were significantly higher than those in the maternal blood (0.56 ± 0.2 vs. 0.12 ± 0.04mM, *p* = 0.003, Mann–Whitney *U* test). In addition, those in the umbilical vein (0.43 ± 0.31, *p* = 0.027, Mann–Whitney *U* test) and artery (0.42 ± 0.26, *p* = 0.045, Mann–Whitney U test) were also higher than those in the maternal blood.

**Figure 1:**
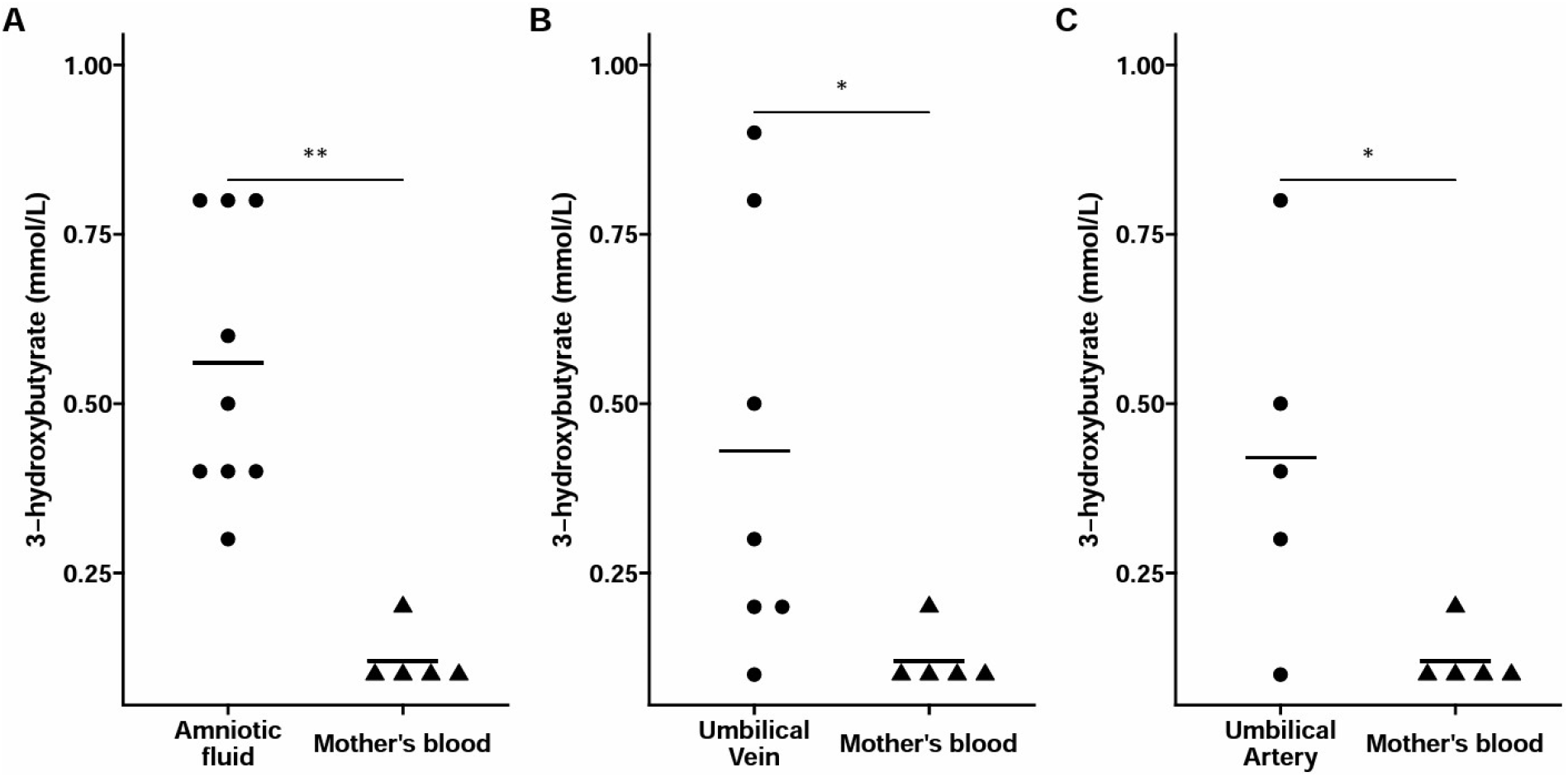
3HB concentrations in various tissues. We compared the 3HB levels of (A) amniotic fluid vs. maternal blood, (B) umbilical vein vs. maternal blood, and (C) artery vs. maternal blood using the Mann–Whitney U test. The circles indicate amniotic fluid or umbilical vein or umbilical artery. The triangles indicate maternal blood. **: *p* < 0.01; ∗ : p < 0.05

### Case

We observed HMGCS2 expression in the placenta right after delivery (Figure 2). Although we had supposed that HMGCS2 would be expressed in the trophoblast of the villi, the expression was significant but it not particularly potent. We discovered that the most potent tissue was the chorionic plate and chorioamniotic membrane, supposedly releasing 3HB into the amniotic fluid.

**Figure 2:**
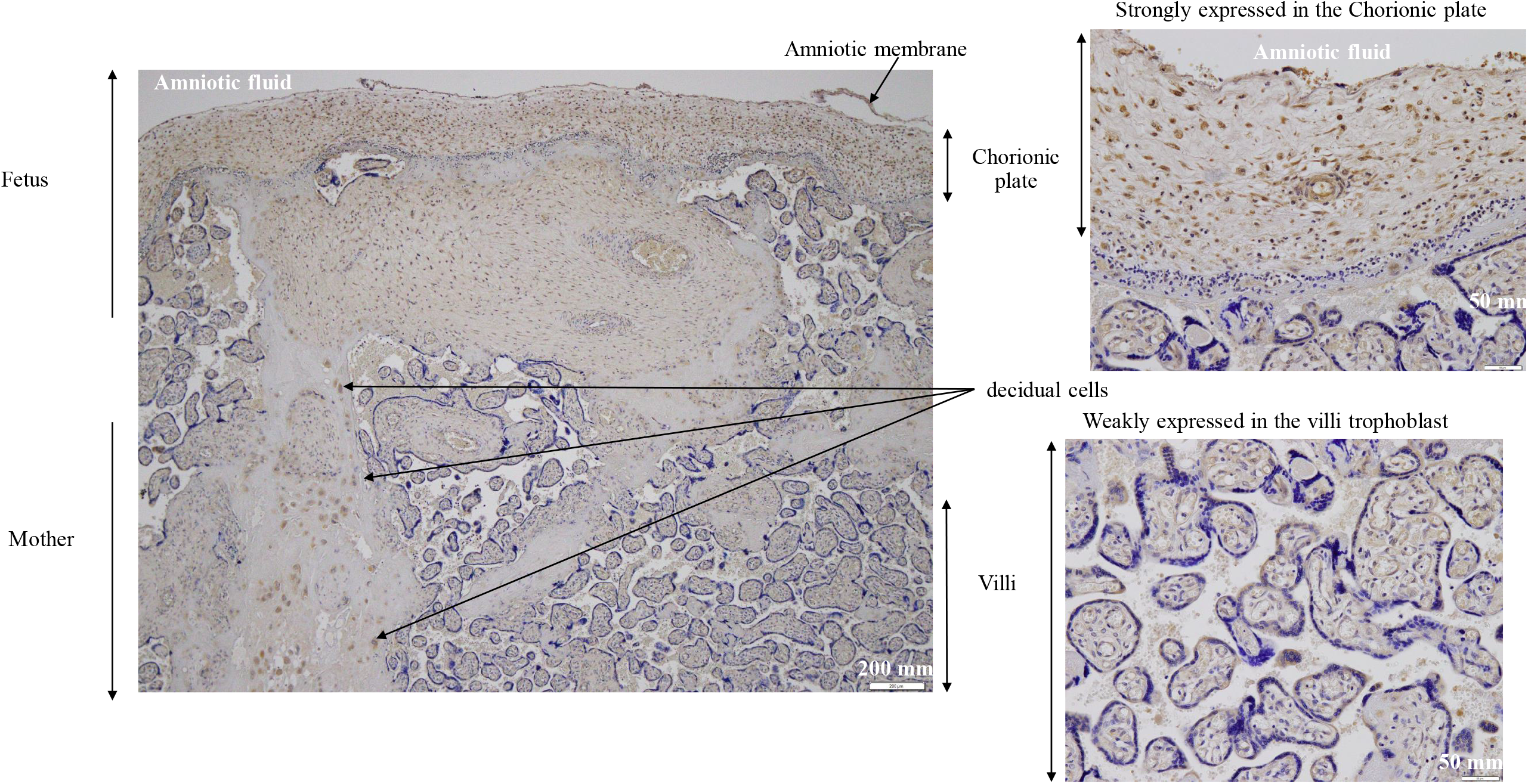
HMGCS2 expression in the placenta. HMGCS2 expression was examined in the human placenta using immunohistochemistry right after delivery. Note that the human placenta displays two 3HB producers, villi and the chorionic plate to supply 3HB to the umbilical vein and amniotic fluid, respectively.

## Discussion

### Two ketone body supply pathways from the placenta

Placenta is one of the most important 3HB suppliers of the human fetus during late pregnancy [11,15]. Since most energy substrates are devoted to the development of the brain during late pregnancy, placental 3HB production should be dedicated to fetal brain development. The placenta exhibits two cell populations that produce 3HB for the fetus: placental villi and the chorionic plate. We present an overview of 3HB flow in Figure 3.

**Figure 3:**
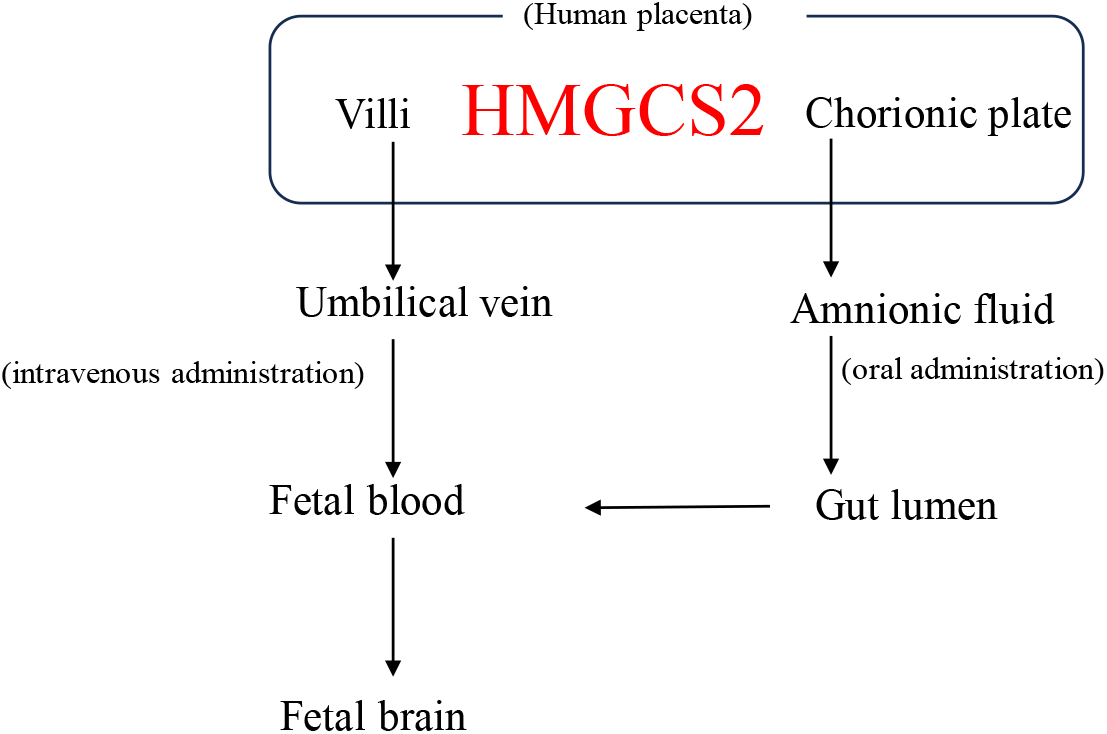
Basic 3HB flow from the placenta to the human fetus. Two potential 3HB delivery pathways exist from the placenta to the human fetus: Pathway 1: Chorionic plate → amniotic fluid → small intestine → fetus Pathway 2: Placental villi trophoblast→ umbilical vein → fetus Note that pathway 1 is a novel finding of this study.

## Data Availability

The data are available from the corresponding author on reasonable request.

## Abbreviations

HMGCS2: 3-hydroxy-3-methylglutaryl-CoA synthase 2
3HB: 3-hydroxybutyrate

## Acknowledgments

The authors thank Mr Takashi Tsujino for creating the graphical design for the Graphical Abstract.

---

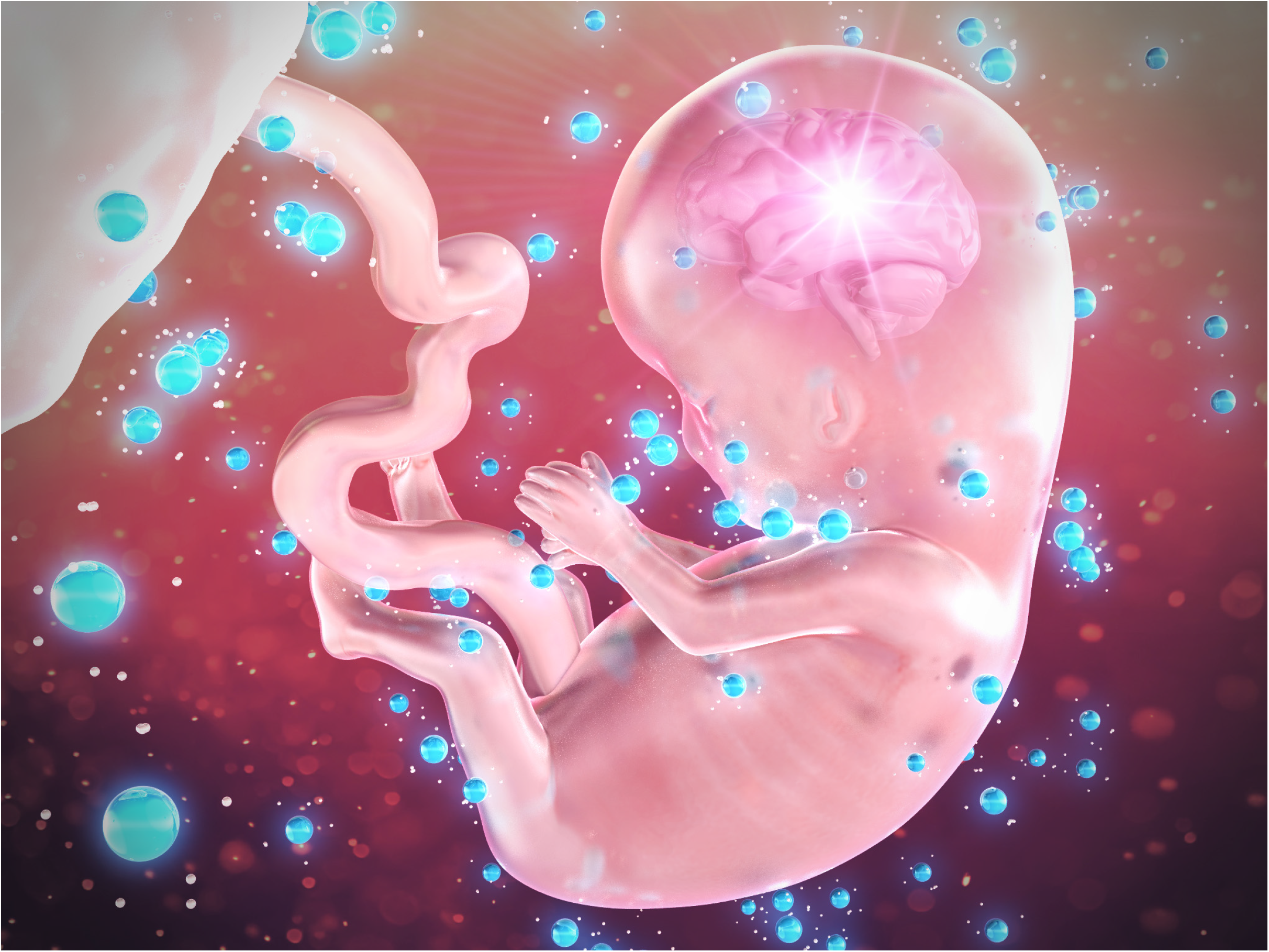

